# The Causal Role of Apolipoprotein B/A1 Ratio in Coronary Heart Disease and Ischemic Stroke: A Mendelian Randomization Study

**DOI:** 10.1101/2025.06.11.25328879

**Authors:** Han Li, Baiju Wang, Hanwen Chen, Yajuan Chen, Yuan Li, Xiaobing Li, Na Wang, Lei Liu, Zihua Song

## Abstract

**Background/Aims:** Although epidemiological analyses have suggested a link between a heightened ratio of apolipoprotein (Apo) B/A1 and atherosclerosis (ATS), the exact causative connection between ApoB/A1 and coronary heart disease (CHD) and ischemic stroke (IS) onset is ambiguous. Accordingly, our objective was to investigate the causal interaction between ApoB/A1 and both CHD and IS.

**Methods:** A Mendelian randomization (MR) approach was applied, utilizing data from two samples to infer causality. Extensive genetic information concerning ApoB/A1 was retrieved from a recent genome-wide association study (GWAS) encompassing 115,082 participants of European ancestry. The SNP datasets associated with CHD and IS were downloaded by accessing the IEU GWAS database. The MR approach was deployed to determine causal impacts and identify pleiotropy.

**Results:** Our results reveal that elevated ApoB/A1 was linked with heightened risks of CHD (OR: 1.008; 95% CI: 1.005–1.010; P < 0.001) and IS (OR: 1.332; 95% CI: 1.112–1.595; P = 0.002).

**Conclusions:** Our findings confirm the causal relationship between a genetic predisposition for an elevated ApoB/A1 and heightened CHD and IS risks. To conclude, ApoB/A1 could be a potential therapeutic target for these two diseases.

## Introduction

Coronary heart disease (CHD) and ischemic stroke (IS) are major drivers of mortality and morbidity globally, representing major cardiovascular diseases that significantly impact public health [1]. Both CHD and IS result from atherosclerosis (ATS), a prevalent condition characterized by lipid accumulation and other blood-derived substances in arterial walls throughout most vascular regions[2]. Cholesterol deposition in the arterial walls is a primary contributor to ATS onset [2]. Apolipoproteins (Apo) are essential for cholesterol transport and metabolism. The ApoB/A1 ratio has become an important biomarker for ATS [3], as it reflects the equilibrium between atherogenic and anti-atherogenic lipoproteins, serving as a crucial cardiovascular risk indicator [4]. Although numerous epidemiological studies have linked ApoB/A1 to both coronary artery disease [5] and stroke risks [6], the causal relationship remains uncertain due to potential confounders and the possibility of reverse causality. Consequently, robust research methodologies are necessary to clarify the ApoB/A1 causal impact on cardiovascular outcomes.

Mendelian randomization (MR) represents a robust approach that leverages genetic variants (GVs) as instrumental variables (IVs) to infer causal links between modifiable risk factors and disease outcomes [7]. Given that GVs are allocated at random during conception, environmental confounders possibly have minimal effect on their associations with disease outcomes [8]. Therefore, MR analyses utilizing GVs as IVs can prevent confounding and reverse causation, enabling stronger causal inferences between exposures and disease risk [8]. As far as we know, no MR study has yet established the causal connection between ApoB/A1 and both CHD and IS risks. The research aims to systematically evaluate the ApoB/A1 causal effect on both CHD and IS risks using MR.

## Methods

### Study design

A two-sample MR design was employed, with three key assumptions underpinning MR studies validity (**Figure 1)**. First, the genetic instruments must exhibit a robust interaction with the exposure. Second, the relation between the genetic instruments and the outcome must remain free from unmeasured confounding. Third, the genetic instruments should directly impact the outcome solely via exposure, without any indirect effects [9].

**Figure 1.**
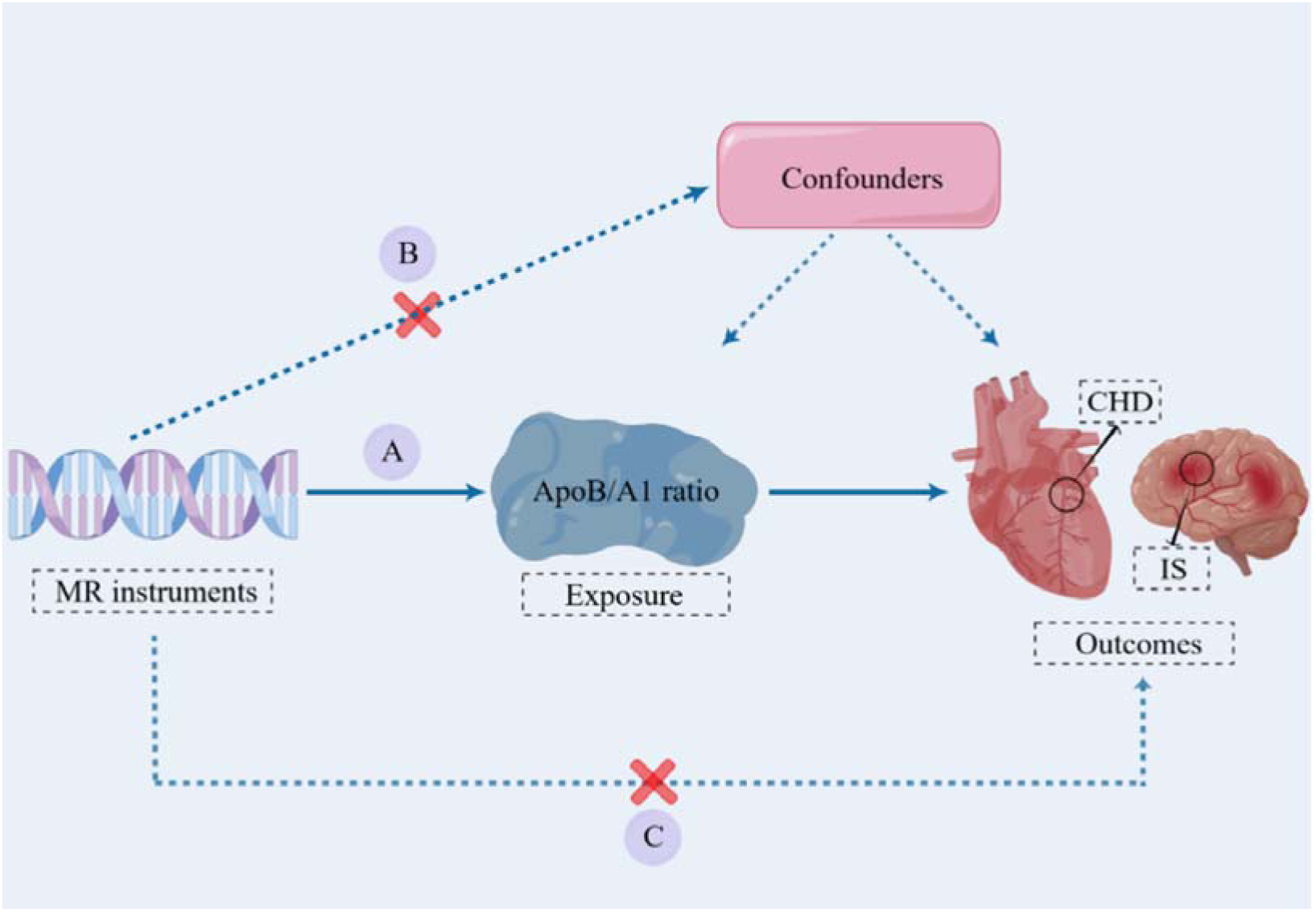
Three key assumptions of MR study. (A) SNPs must be associated with ApoB/A1 ratio; (B) SNPs must be independent of confounders; (C) SNPs should not be directly associated with outcomes in patients. MR, Mendelian randomization; SNP, single nucleotide polymorphism; ApoB/A1 ratio: ratio of Apolipoprotein B to Apolipoprotein A1; CHD, coronary heart disease; IS, ischemic stroke.

### Data sources

The exposure factor, plasma ApoB/A1, was selected from the IEU GWAS database (ebi-a-GCST90092810), comprising 115,082 serum samples [10, 11]. Exposure variable-related single nucleotide polymorphisms (SNPs) were identified, and summary statistics (effect size, standard error, and P-value) were documented. A total of 115,082 relevant SNPs were extracted and included in a meta-analysis.

All genetic instruments demonstrated an association with ApoB/A1 at the genome-wide significance level (p < 5E−8). To guarantee the independence of the selected SNPs, linkage disequilibrium (LD) among ApoB/A1-related SNPs was evaluated via the PLINK clumping method, which relies on the European reference panel from the 1,000 Genomes project. The SNPs were considered independent if they had an LD r^2^ < 0.01 within a 10 Kb window, and these independent SNPs were subsequently utilized as IVs in this investigation. Ultimately, 72 significant SNPs linked to ApoB/A1 were derived through this meta-analysis. **Table 1** lists a comprehensive list of these genetic instruments.

**Table 1.**
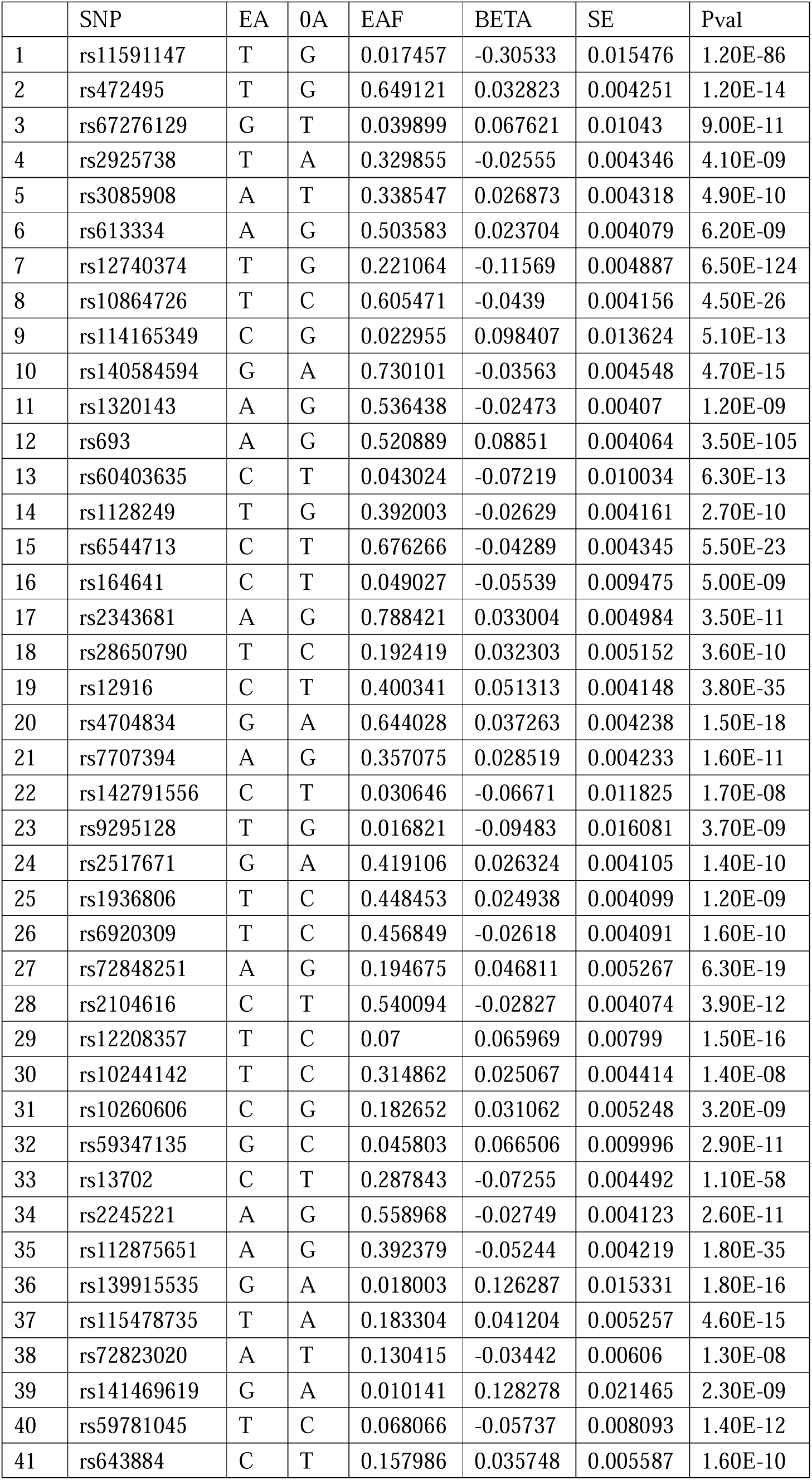
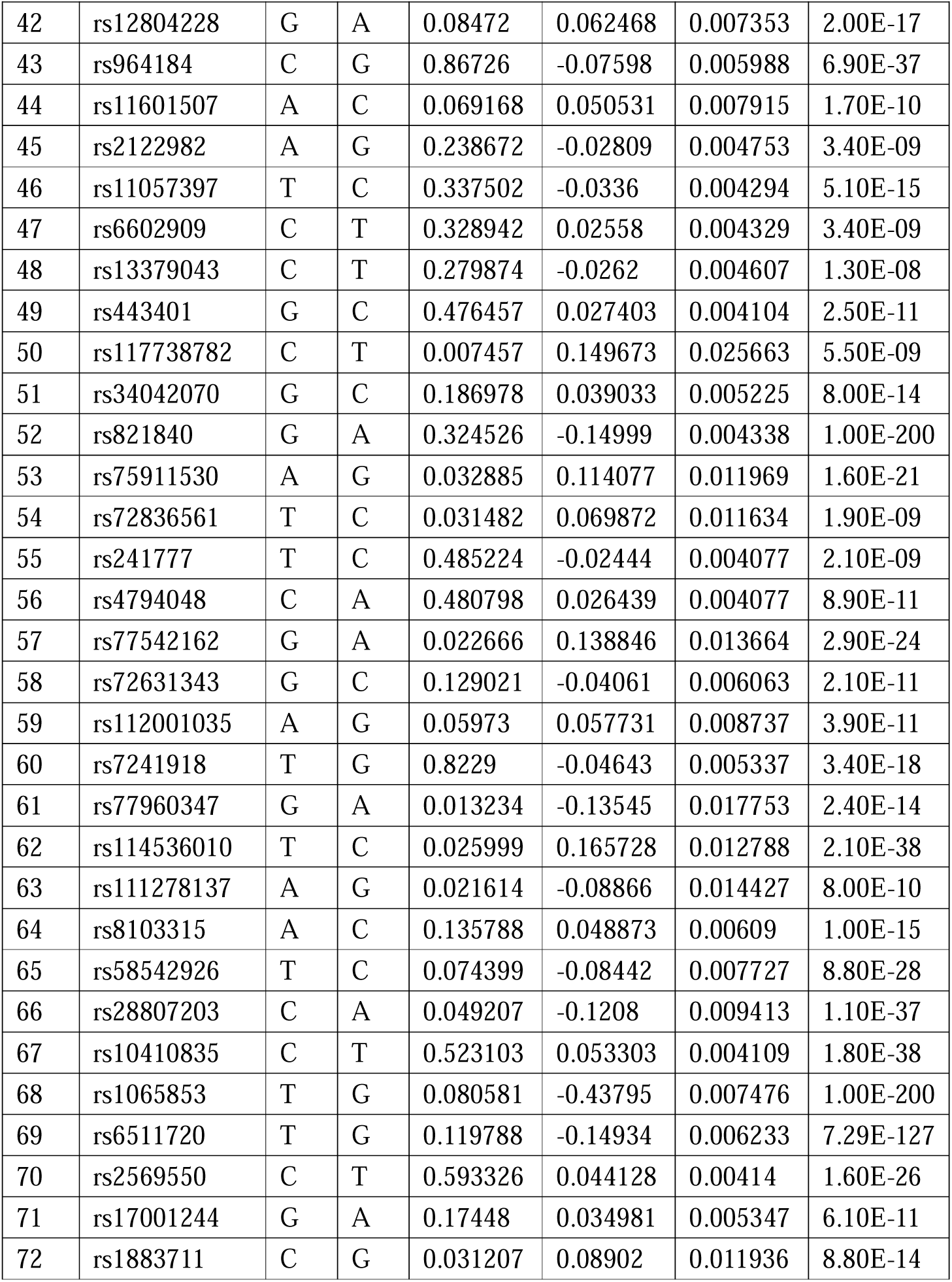
The characteristics of 72 SNPs and their associations with ApoB/A1 ratio.

|  | SNP | EA | OA | EAF | BETA | SE | Pval |
| --- | --- | --- | --- | --- | --- | --- | --- |
| 1 | rs11591147 | T | G | 0.017457 | -0.30533 | 0.015476 | 1.20E-86 |
| 2 | rs472495 | T | G | 0.649121 | 0.032823 | 0.004251 | 1.20E-14 |
| 3 | rs67276129 | G | T | 0.039899 | 0.067621 | 0.01043 | 9.00E-11 |
| 4 | rs2925738 | T | A | 0.329855 | -0.02555 | 0.004346 | 4.10E-09 |
| 5 | rs3085908 | A | T | 0.338547 | 0.026873 | 0.004318 | 4.90E-10 |
| 6 | rs613334 | A | G | 0.503583 | 0.023704 | 0.004079 | 6.20E-09 |
| 7 | rs12740374 | T | G | 0.221064 | -0.11569 | 0.004887 | 6.50E-124 |
| 8 | rs10864726 | T | C | 0.605471 | -0.0439 | 0.004156 | 4.50E-26 |
| 9 | rs114165349 | C | G | 0.022955 | 0.098407 | 0.013624 | 5.10E-13 |
| 10 | rs140584594 | G | A | 0.730101 | -0.03563 | 0.004548 | 4.70E-15 |
| 11 | rs1320143 | A | G | 0.536438 | -0.02473 | 0.00407 | 1.20E-09 |
| 12 | rs693 | A | G | 0.520889 | 0.08851 | 0.004064 | 3.50E-105 |
| 13 | rs60403635 | C | T | 0.043024 | -0.07219 | 0.010034 | 6.30E-13 |
| 14 | rs1128249 | T | G | 0.392003 | -0.02629 | 0.004161 | 2.70E-10 |
| 15 | rs6544713 | C | T | 0.676266 | -0.04289 | 0.004345 | 5.50E-23 |
| 16 | rs164641 | C | T | 0.049027 | -0.05539 | 0.009475 | 5.00E-09 |
| 17 | rs2343681 | A | G | 0.788421 | 0.033004 | 0.004984 | 3.50E-11 |
| 18 | rs28650790 | T | C | 0.192419 | 0.032303 | 0.005152 | 3.60E-10 |
| 19 | rs12916 | C | T | 0.400341 | 0.051313 | 0.004148 | 3.80E-35 |
| 20 | rs4704834 | G | A | 0.644028 | 0.037263 | 0.004238 | 1.50E-18 |
| 21 | rs7707394 | A | G | 0.357075 | 0.028519 | 0.004233 | 1.60E-11 |
| 22 | rs142791556 | C | T | 0.030646 | -0.06671 | 0.011825 | 1.70E-08 |
| 23 | rs9295128 | T | G | 0.016821 | -0.09483 | 0.016081 | 3.70E-09 |
| 24 | rs2517671 | G | A | 0.419106 | 0.026324 | 0.004105 | 1.40E-10 |
| 25 | rs1936806 | T | C | 0.448453 | 0.024938 | 0.004099 | 1.20E-09 |
| 26 | rs6920309 | T | C | 0.456849 | -0.02618 | 0.004091 | 1.60E-10 |
| 27 | rs72848251 | A | G | 0.194675 | 0.046811 | 0.005267 | 6.30E-19 |
| 28 | rs2104616 | C | T | 0.540094 | -0.02827 | 0.004074 | 3.90E-12 |
| 29 | rs12208357 | T | C | 0.07 | 0.065969 | 0.00799 | 1.50E-16 |
| 30 | rs10244142 | T | C | 0.314862 | 0.025067 | 0.004414 | 1.40E-08 |
| 31 | rs10260606 | C | G | 0.182652 | 0.031062 | 0.005248 | 3.20E-09 |
| 32 | rs59347135 | G | C | 0.045803 | 0.066506 | 0.009996 | 2.90E-11 |
| 33 | rs13702 | C | T | 0.287843 | -0.07255 | 0.004492 | 1.10E-58 |
| 34 | rs2245221 | A | G | 0.558968 | -0.02749 | 0.004123 | 2.60E-11 |
| 35 | rs112875651 | A | G | 0.392379 | -0.05244 | 0.004219 | 1.80E-35 |
| 36 | rs139915535 | G | A | 0.018003 | 0.126287 | 0.015331 | 1.80E-16 |
| 37 | rs115478735 | T | A | 0.183304 | 0.041204 | 0.005257 | 4.60E-15 |
| 38 | rs72823020 | A | T | 0.130415 | -0.03442 | 0.00606 | 1.30E-08 |
| 39 | rs141469619 | G | A | 0.010141 | 0.128278 | 0.021465 | 2.30E-09 |
| 40 | rs59781045 | T | C | 0.068066 | -0.05737 | 0.008093 | 1.40E-12 |
| 41 | rs643884 | C | T | 0.157986 | 0.035748 | 0.005587 | 1.60E-10 |
| 42 | rs12804228 | G | A | 0.08472 | 0.062468 | 0.007353 | 2.00E-17 |
| 43 | rs964184 | C | G | 0.86726 | -0.07598 | 0.005988 | 6.90E-37 |
| 44 | rs11601507 | A | C | 0.069168 | 0.050531 | 0.007915 | 1.70E-10 |
| 45 | rs2122982 | A | G | 0.238672 | -0.02809 | 0.004753 | 3.40E-09 |
| 46 | rs11057397 | T | C | 0.337502 | -0.0336 | 0.004294 | 5.10E-15 |
| 47 | rs6602909 | C | T | 0.328942 | 0.02558 | 0.004329 | 3.40E-09 |
| 48 | rs13379043 | C | T | 0.279874 | -0.0262 | 0.004607 | 1.30E-08 |
| 49 | rs443401 | G | C | 0.476457 | 0.027403 | 0.004104 | 2.50E-11 |
| 50 | rs117738782 | C | T | 0.007457 | 0.149673 | 0.025663 | 5.50E-09 |
| 51 | rs34042070 | G | C | 0.186978 | 0.039033 | 0.005225 | 8.00E-14 |
| 52 | rs821840 | G | A | 0.324526 | -0.14999 | 0.004338 | 1.00E-200 |
| 53 | rs75911530 | A | G | 0.032885 | 0.114077 | 0.011969 | 1.60E-21 |
| 54 | rs72836561 | T | C | 0.031482 | 0.069872 | 0.011634 | 1.90E-09 |
| 55 | rs241777 | T | C | 0.485224 | -0.02444 | 0.004077 | 2.10E-09 |
| 56 | rs4794048 | C | A | 0.480798 | 0.026439 | 0.004077 | 8.90E-11 |
| 57 | rs77542162 | G | A | 0.022666 | 0.138846 | 0.013664 | 2.90E-24 |
| 58 | rs72631343 | G | C | 0.129021 | -0.04061 | 0.006063 | 2.10E-11 |
| 59 | rs112001035 | A | G | 0.05973 | 0.057731 | 0.008737 | 3.90E-11 |
| 60 | rs7241918 | T | G | 0.8229 | -0.04643 | 0.005337 | 3.40E-18 |
| 61 | rs77960347 | G | A | 0.013234 | -0.13545 | 0.017753 | 2.40E-14 |
| 62 | rs114536010 | T | C | 0.025999 | 0.165728 | 0.012788 | 2.10E-38 |
| 63 | rs111278137 | A | G | 0.021614 | -0.08866 | 0.014427 | 8.00E-10 |
| 64 | rs8103315 | A | C | 0.135788 | 0.048873 | 0.00609 | 1.00E-15 |
| 65 | rs58542926 | T | C | 0.074399 | -0.08442 | 0.007727 | 8.80E-28 |
| 66 | rs28807203 | C | A | 0.049207 | -0.1208 | 0.009413 | 1.10E-37 |
| 67 | rs10410835 | C | T | 0.523103 | 0.053303 | 0.004109 | 1.80E-38 |
| 68 | rs1065853 | T | G | 0.080581 | -0.43795 | 0.007476 | 1.00E-200 |
| 69 | rs6511720 | T | G | 0.119788 | -0.14934 | 0.006233 | 7.29E-127 |
| 70 | rs2569550 | C | T | 0.593326 | 0.044128 | 0.00414 | 1.60E-26 |
| 71 | rs17001244 | G | A | 0.17448 | 0.034981 | 0.005347 | 6.10E-11 |
| 72 | rs1883711 | C | G | 0.031207 | 0.08902 | 0.011936 | 8.80E-14 |

The SNP datasets related to CHD and IS were acquired from the IEU GWAS database (v8.0, April 2024) [12], and we followed the IEU GWAS database terms of use and acknowledged the database team for their support. Notably, all the data we applied were derived from participants of European ancestry. Since we only used publicly available datasets without direct participant involvement, ethics approval was not required.

### IV Extraction

To satisfy the Assumption 1 relevance criterion, we chose independent SNPs (R^2^ < 0.001, 10,000 kb threshold) from the original set of 72 SNPs. These SNPs exhibited genome-wide significant associations with plasma ApoB/A1 (p < 5E−8) and were designated as potential IVs.

Furthermore, to verify that the IVs had sufficient power to discern the exposure causal effect on the outcome, F-statistics for these potential IVs were computed via an online platform (https://sb452.shinyapps). An F-statistic > 10 indicates that the IVs are robust and appropriate for analysis. To meet Assumptions 2–3, specifically the independence and exclusion restriction assumptions, potential confounders for plasma ApoB/A1, CHD, and IS were initially identified through meta-analysis. Subsequently, each candidate IV was screened using the PhenoScanner database to detect associations with these confounders. Candidate IVs with significant interconnections to the outcome/confounder (p < 5E−8) were excluded from subsequent analysis, advancing the remaining IVs to the MR analysis preliminary phase. Due to the ambiguous biological roles of several GVs, the MR-PRESSO method was applied to define and exclude potential outliers before conducting each MR analysis.

### MR Analysis

For the assessment of the causal relations between ApoB/A1 and both CHD and IS risks, we deployed the inverse variance-weighted MR (IVW-MR) that combines Wald ratios from individual SNPs via meta-analysis, providing an in-depth estimation of the exposure’s effect on the outcome. The IVW-MR can be performed with a fixed- or random-effects model, with the latter applied in this study when significant heterogeneity (p < 0.05) was detected. To validate and supplement the IVW-MR results, we employed additional techniques: MR-Egger, weighted median, penalized weighted median, and maximum likelihood estimation. A Bonferroni correction for multiple comparisons was deployed, with a significance threshold of 0.0125 (0.05/4). Analyses were carried out with R software v4.1.2, utilizing the two-sample MR package.

### Sensitivity Analysis

To assess our MR estimates robustness, several sensitivity analyses were performed. First, Cochran’s Q test was employed to assess heterogeneity across GVs. Subsequently, we utilized the MR-Egger intercept and MR-PRESSO global tests to explore the probable influence of pleiotropy on the outcomes. Lastly, a leave-one-out (LOO) analysis was performed to assess whether excluding any individual SNP possessed a substantial effect on the overall estimate.

## Results

### SNP selection and validation

To meet the requirements of MR studies and ensure the validity of IVs, we preprocessed the obtained exposure SNPs by excluding those directly associated with the outcome variables to avoid violating the independence assumption. Using the LD clumping method with a threshold of r² < 0.01, we ensured the independence of the selected SNPs. Ultimately, 72 SNPs were classified as IVs in our study. Among them, 39 SNPs were linked to an elevation in plasma ApoB/A1, whereas 33 SNPs were related to a decrease in genetically predicted plasma ApoB/A1. In particular, all selected SNPs possessed F-statistics > 10, implying minimal bias risk from weak Ivs. **Table 1** summarizes further details on the IVs and their associations with each outcome.

### MR estimates

The IVW analysis results indicated that ApoB/A1 serves as a risk factor for both CHD and IS (**Tables 2– 3**). Among them, the analysis yielded an odds ratio (OR) of 1.008 (95% CI: 1.005-1.010; P < 0.001) for CHD and 1.332 (95% CI: 1.112-1.595; P = 0.002) for IS. While some MR methods failed to achieve statistical significance, their results consistently pointed in the same direction. Furthermore, the forest (**Figure 2**) and scatter plots (**Figure 3**) underscore the positive connection between ApoB/A1 and escalated CHD and IS risks.

**Figure 2.**
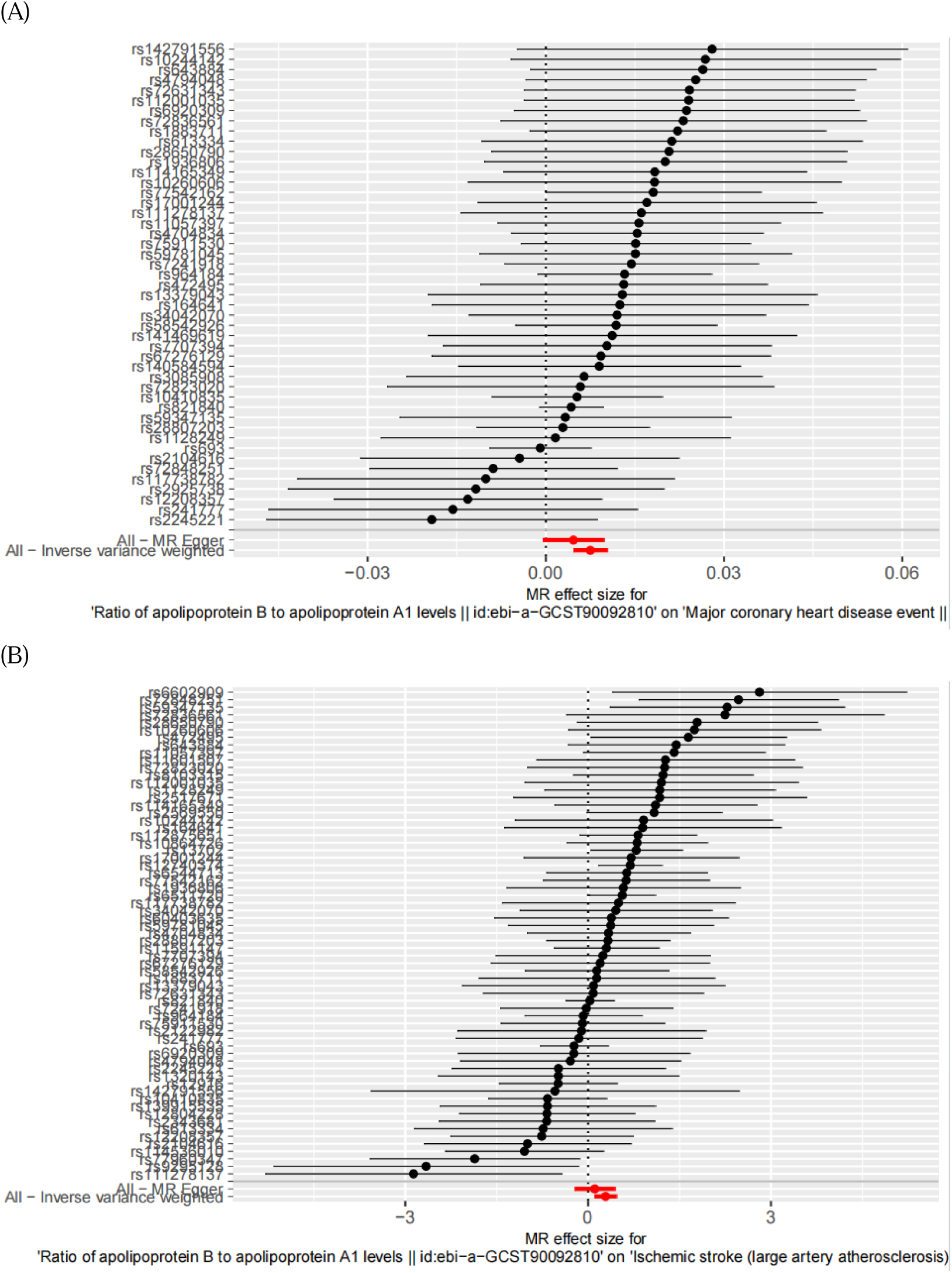
Analysis of the SNP effects on the development of CHD and IS. (A) ApoB/A1 ratio-CHD; (B) ApoB/A1 ratio-IS. In the forest map, each black dot represents a single SNP as instrumental variable and the red dot shows the use of IVW results for all SNPs. ApoB/A1 ratio: ratio of Apolipoprotein B to Apolipoprotein A1; CHD, coronary heart disease; IS, ischemic stroke; SNP, single nucleotide polymorphism; IVW, inverse variance weighted.

**Figure 3.**
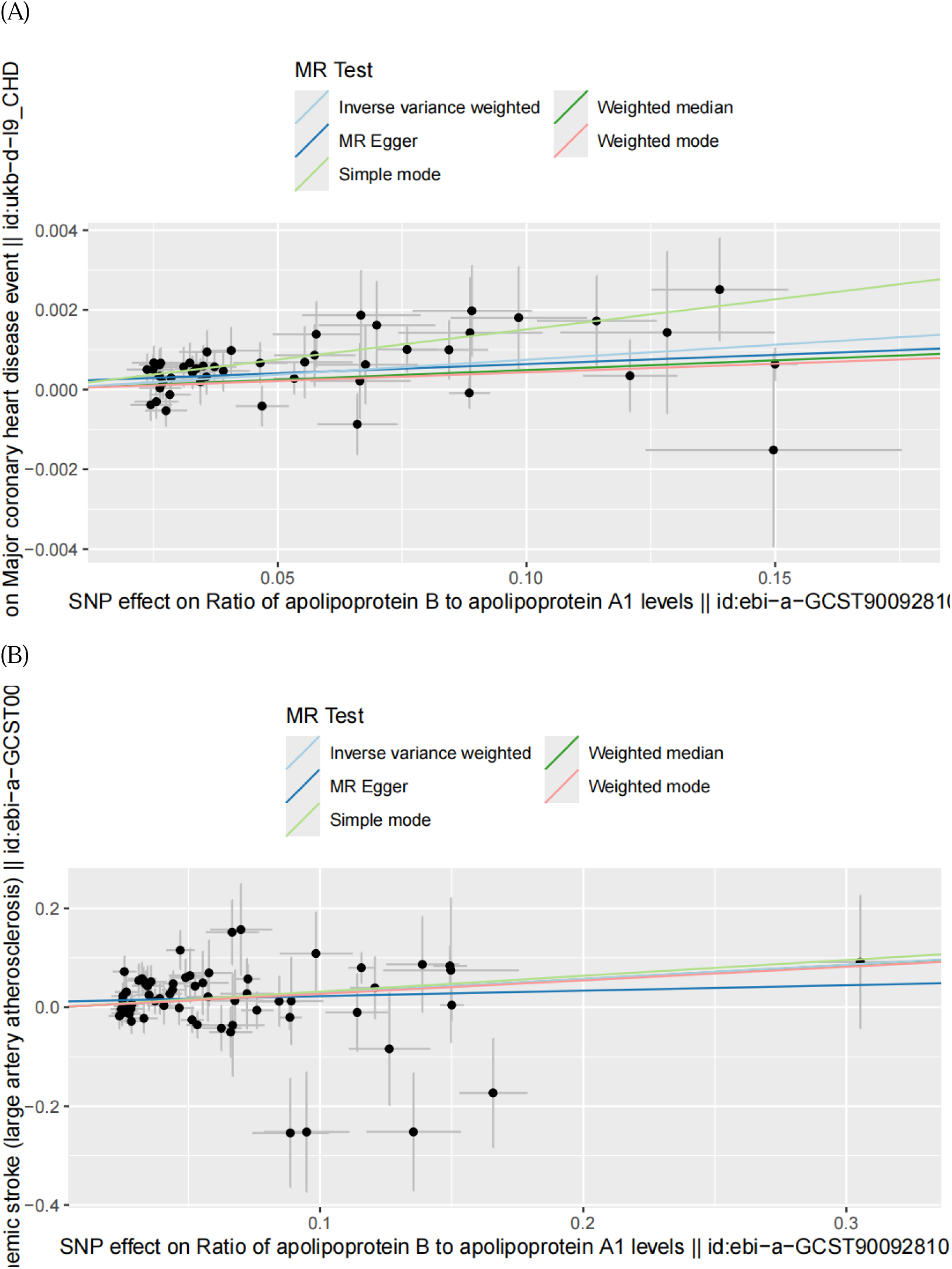
Scatter plots of the estimated SNP effects on ApoB/A1 ratio (x-axis) plotted against the estimated SNPs effects on CHD and IS (y-axis). (A) ApoB/A1 ratio-CHD; (B) ApoB/A1 ratio-IS. The slope of the line corresponds to a causal estimate using a different method. ApoB/A1 ratio: ratio of Apolipoprotein B to Apolipoprotein A1; CHD, coronary heart disease; IS, ischemic stroke.

**Table 2.** Association of ApoB/A1 ratio levels and risk of CHD in patients using different methods.

| Method | OR | OR_lci95 | OR_uci95 | Pval |
| --- | --- | --- | --- | --- |
| MR Egger | 1.0046552 | 0.999501 | 1.009836 | 0.08351425 |
| Weighted median | 1.0049091 | 1.000439 | 1.0094 | 0.03133307 |
| Inverse variance weighted | 1.0075229 | 1.004655 | 1.010399 | 2.55E-07 |
| Simple mode | 1.0152142 | 1.003584 | 1.026979 | 0.01351933 |
| Weighted mode | 1.0043278 | 0.999767 | 1.008909 | 0.06932352 |

**Table 3.** Association of ApoB/A1 ratio levels and risk of IS in patients using different methods.

| Method | OR | OR_lci95 | OR_uci95 | Pval |
| --- | --- | --- | --- | --- |
| MR Egger | 1.116928 | 0.804248 | 1.551173 | 0.511704 |
| Weighted median | 1.329251 | 1.0293 | 1.716613 | 0.029159 |
| Inverse variance weighted | 1.331572 | 1.111805 | 1.59478 | 0.00186 |
| Simple mode | 1.374708 | 0.773479 | 2.443277 | 0.282167 |
| Weighted mode | 1.31276 | 0.947366 | 1.819085 | 0.106934 |

### Sensitivity analyses

The LOO results consistently verified the direction and magnitude of the connection between increased plasma ApoB/A1 and both CHD and IS risks (**Figure 4**). In addition, MR analysis revealed no heterogeneity between ApoB/A1 and CHD risk, while heterogeneity with IS risk was observed (**Table 4**). Furthermore, the MR-Egger intercept test further indicated horizontal pleiotropy absence, with all p > 0.05 (**Table 5**).

**Figure 4.**
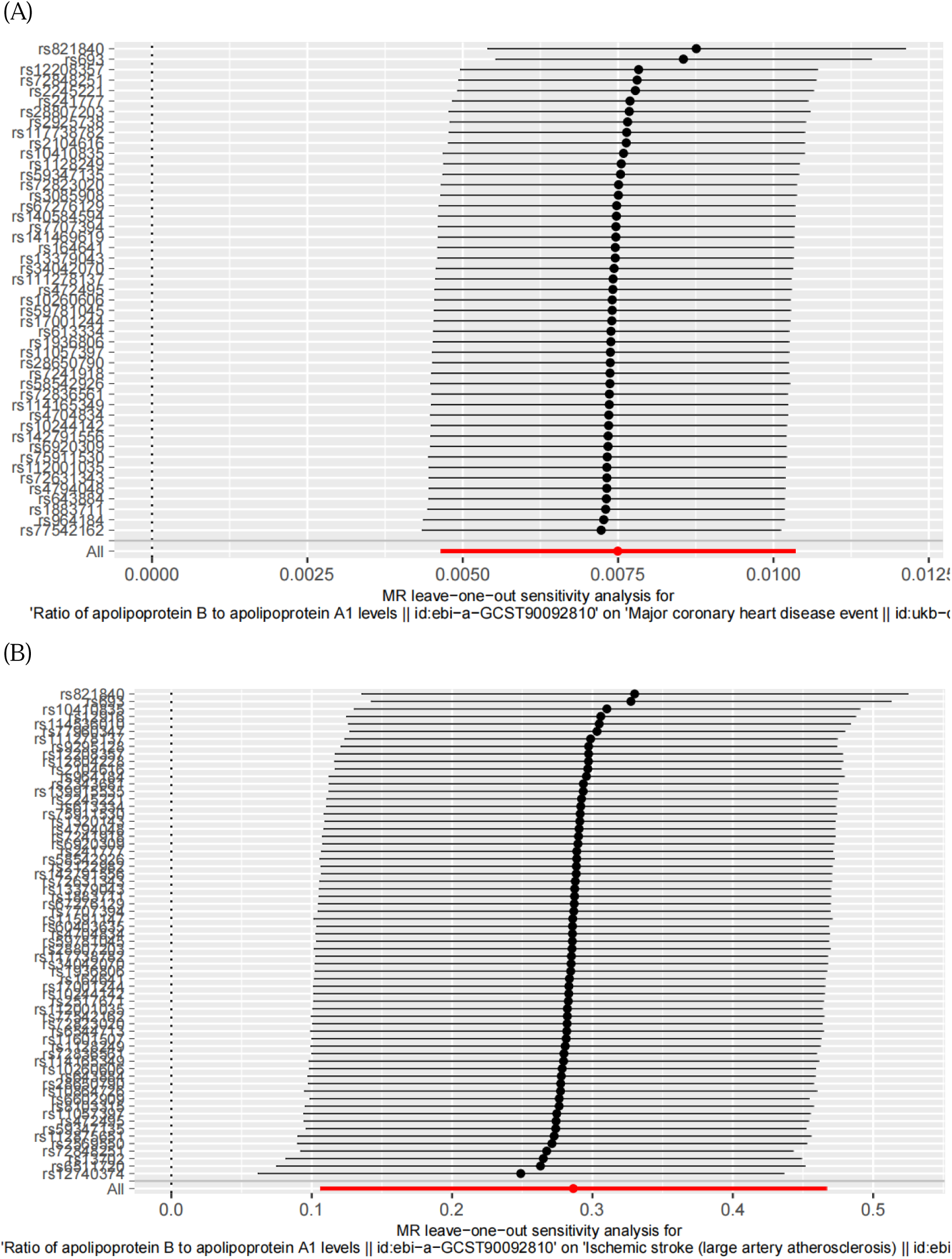
Sensitivity analyses using the leave-one-out approach on the association of ApoB/A1 ratio with CHD and IS. (A) ApoB/A1 ratio-CHD; (B) ApoB/A1 ratio-IS. Each black dot represents an IVW method to estimate the causal effect of the exposures on the CAD and IS. The presence of a particular SNP causing a significant change in the overall results is not excluded. ApoB/A1 ratio: ratio of Apolipoprotein B to Apolipoprotein A1; CHD, coronary heart disease; IS, ischemic stroke; SNP, single nucleotide polymorphism; IVW, inverse variance weighted.

**Table 4.** The results of heterogeneity analysis.

| Exposures | Outcome | method | Q | Pval |
| --- | --- | --- | --- | --- |
| ApoB/A1 ratio | CHD | MR Egger | 40.078801 | 0.680089766 |
| ApoB/A1 ratio | CHD | Inverse variance weighted | 41.7816641 | 0.649467619 |
| ApoB/A1 ratio | IS | MR Egger | 87.9966581 | 0.020490856 |
| ApoB/A1 ratio | IS | Inverse variance weighted | 90.1891895 | 0.017205873 |

**Table 5.** MR-Egger regression intercept.

| Exposure | Outcome | Intercept | SE | Pval |
| --- | --- | --- | --- | --- |
| ApoB/A1 ratio | CHD | 0.000176 | 0.00014 | 0.198548 |
| ApoB/A1 ratio | IS | 0.011493 | 0.00917 | 0.214879 |

## Discussion

Currently, the genetic connection between ApoB/A1 and both CHD and IS risks is unclear. Therefore, we are the first to conduct an MR analysis aimed at elucidating the genetic causal link between ApoB/A1 and the risks of CHD and IS. The results reveal that for each standard deviation elevation in ApoB/A1, the risks of CHD rises by 0.8%, and that of IS escalates by 33.2%. Importantly, the MR analysis showcased no evidence of horizontal pleiotropy between ApoB/A1 and both CHD and IS risks. Additionally, we found no heterogeneity between ApoB/A1 and CHD risk, while heterogeneity was present with IS risk. However, this heterogeneity does not affect the IVW results, rendering our conclusions robust. Moreover, the robustness and consistency of our findings were validated through various analytical techniques. Together, these results underscore a strong interaction between elevated ApoB/A1 and both CHD and IS risk, emphasizing the potential of ApoB/A1 as a crucial diagnostic and therapeutic target for CHD and IS.

Our findings are supported by an abundance of pathophysiological investigations. Presently, ApoB/A1 is pivotal in ATS pathophysiology by indicating the balance between atherogenic and protective lipoproteins [13]. The ApoB, associated with low-density lipoprotein (LDL) and very LDL (VLDL), facilitates cholesterol transport and deposition into the arterial walls [14]. This deposition initiates the formation of fatty streaks and atherosclerotic plaques, which are characterized by lipid accumulation, inflammation, foam cell formation, and potential calcification [14, 15]. These processes contribute to the thickening and hardening of arterial walls, ultimately narrowing arteries and restricting blood flow [8, 14]. Conversely, ApoA1 is the main component of high-density lipoprotein (HDL), which is crucial in reverse cholesterol transport [16]. The ApoA1 interacts with ATP-binding cassette transporters on cell membranes to promote cholesterol efflux from peripheral tissues, including the arterial wall [17]. This mechanism is vital for maintaining lipid homeostasis and preventing the buildup of cholesterol in the arteries. A higher ApoB/A1 signifies an imbalance where atherogenic particles predominate over protective HDL particles [18]. This imbalance enhances lipid deposition within the arterial walls and reduces the efficiency of cholesterol removal, promoting the development and instability of atherosclerotic plaques [19]. Furthermore, Yusuf et al. have highlighted that the ratio exhibits a superior capacity to predict cardiovascular risk when compared to ApoB or ApoA1 individually or any other cholesterol-related metrics [20]. They underscore the crucial role of ApoB/A1 in ATS, though additional research is required to uncover the underlying mechanisms.

Our results align with the observations documented in several prior studies. O’Donnell et al. recruited 26,919 participants from 32 countries, including 13,447 cases of first acute stroke and 13,472 controls, manifesting a robust link between ApoB/A1 and IS across all regions, with a population-attributable risk ranging from 24.8% in Western Europe, North America, and Australia to 67.6% in Southeast Asia [21]. A clinical investigation by Jung et al., encompassing 1,401 participants, has revealed that an elevated serum ApoB/A1 is independently related to significant coronary ATS and noncalcified plaques, the latter being a characteristic feature of early-stage ATS - this finding suggests the serum ApoB/A1 may be particularly valuable in determining incipient atherosclerotic lesions [22]. Furthermore, Yusuf et al. conducted a standardized case-control study on acute myocardial infarction spanning 52 countries. The results indicated that ApoB/A1 is the most critical risk factor in all geographic regions. Consequently, significantly altering its population distribution is essential for mitigating myocardial infarction rates worldwide [20]. Khan et al. conducted a study involving 1,582 case-control pairs across 32 countries to investigate frequent risk factors for stroke in pateints aged less than 45 years. The results indicated that, similar to older adults, hypertension, excessive alcohol consumption, smoking, ApoB/A1, psychosocial stress, and obesity are significant risk factors in the younger population [23]. These observations underscore the importance of ApoB/A1 in atherosclerotic disease. While these observational investigations offer invaluable insights, they fall short of definitively establishing causality because of the possibility of confounding variables and reverse causation bias [24]. Consequently, the direct causal connection between ApoB/A1 and both CHD and IS risks remains ambiguous. Our study employs MR to reveal a significant genetic link between ApoB/A1 and both CHD and IS risks, thereby offering evidence for a direct causal relation in the European population.

Notably, our survey is limited; first, the exclusive utilization of European samples in the GWAS may constrain the result’s generalizability to other ethnic populations [25]. Second, the presence of pleiotropy introduces challenges in fully eliminating bias from our estimates. Although SNPs linked to confounding factors were excluded, residual effects may still remain. Furthermore, the study is limited to examining the genetic connection between ApoB/A1 and CHD and IS without accounting for environmental influences. Finally, despite our study providing evidence for the interaction between ApoB/A1 and CHD and IS, the molecular mechanisms behind ApoB/A1 in these diseases remain unclear. Future studies should include diverse ethnic groups, account for both genetic and environmental factors, and involve further *in vitro* and *in vivo* investigations to elucidate the specific molecular mechanisms.

## Conclusion

Our research reveals a substantial genetic correlation between ApoB/A1 and both CHD and IS risks, indicating the potential of ApoB/A1 to be a valuable therapeutic target for preventing and treating these conditions.

## Data availability

Data are provided within the manuscript.

## Data Availability

Data are provided within the manuscript.

## Authors’ Contributions

LL and SZH conceptualized the study. LH and WBJ were involved in the study design, performed statistical analyses, and wrote the manuscript. CHW, CYJ, and LY contributed to the study design and assisted in manuscript preparation. LXB and WN played a role in acquiring genetic association data. All authors reviewed and approved the final manuscript for publication.

## Acknowledgments

Funded by the Research Fund for Academician Lin He New Medicine (no. JYHL2021FMS17).

## References

[1] Tsao, C. W., Aday, A. W., Almarzooq, Z. I., Anderson, C. A. M., Arora, P., Avery, C. L., Baker-Smith, C. M., Beaton, A. Z., Boehme, A. K., Buxton, A. E., Commodore-Mensah, Y., Elkind, M. S. V., Evenson, K. R., Eze-Nliam, C., Fugar, S., Generoso, G., Heard, D. G., Hiremath, S., Ho, J. E., Kalani, R., … American Heart Association Council on Epidemiology and Prevention Statistics Committee and Stroke Statistics Subcommittee (2023). Heart Disease and Stroke Statistics-2023 Update: A Report From the American Heart Association. Circulation, *147*(8), e93–e621. 10.1161/CIR.0000000000001123

[2] Conforto, A. B., Leite, C.daC., Nomura, C. H., Bor-Seng-Shu, E., & Santos, R. D. (2013). Is there a consistent association between coronary heart disease and ischemic stroke caused by intracranial atherosclerosis?. Arquivos de neuro-psiquiatria, 71(5), 320–326. 10.1590/0004-282x20130028

[3] Marcovina, S., & Packard, C. J. (2006). Measurement and meaning of apolipoprotein AI and apolipoprotein B plasma levels. Journal of internal medicine, 259(5), 437–446. 10.1111/j.1365-2796.2006.01648.x

[4] Walldius, G., Jungner, I., Holme, I., Aastveit, A. H., Kolar, W., & Steiner, E. (2001). High apolipoprotein B, low apolipoprotein A-I, and improvement in the prediction of fatal myocardial infarction (AMORIS study): a prospective study. *Lancet (London*, England), 358(9298), 2026–2033. 10.1016/S0140-6736(01)07098-2

[5] Hua, R., Li, Y., Li, W., Wei, Z., Yuan, Z., & Zhou, J. (2021). Apolipoprotein B/A1 Ratio Is Associated with Severity of Coronary Artery Stenosis in CAD Patients but Not in Non-CAD Patients Undergoing Percutaneous Coronary Intervention. Disease markers, 2021, 8959019. 10.1155/2021/8959019

[6] Dong, H., Chen, W., Wang, X., Pi, F., Wu, Y., Pang, S., Xie, Y., Xia, F., & Zhang, Q. (2015). Apolipoprotein A1, B levels, and their ratio and the risk of a first stroke: a meta-analysis and case-control study. Metabolic brain disease, 30(6), 1319–1330. 10.1007/s11011-015-9732-7

[7] Wu, E., Ni, J. T., Xie, T., & Tao, L. (2022). Noncausal effects of genetic predicted depression and colorectal cancer risk: A Mendelian randomization study. Medicine, 101(34), e30177. 10.1097/MD.0000000000030177

[8] Zheng, P. F., Rong, J. J., Zheng, Z. F., Liu, Z. Y., Pan, H. W., & Liu, P. (2023). Investigating the causal effect of Dickkopf-1 on coronary artery disease and ischemic stroke: a Mendelian randomization study. Aging, 15(18), 9797–9808. 10.18632/aging.205050

[9] Wang, B., Li, H., Wang, N., Li, Y., Song, Z., Chen, Y., Li, X., Liu, L., & Chen, H. (2025). The impact of homocysteine on patients with diabetic nephropathy: a mendelian randomization study. Acta diabetologica, 62(1), 123–130. 10.1007/s00592-024-02343-9

[10] Gkatzionis, A., Burgess, S., & Newcombe, P. J. (2023). Statistical methods for cis-Mendelian randomization with two-sample summary-level data. Genetic epidemiology, 47(1), 3–25. 10.1002/gepi.22506

[11] Richardson, T. G., Leyden, G. M., Wang, Q., Bell, J. A., Elsworth, B., Davey Smith, G., & Holmes, M. V. (2022). Characterising metabolomic signatures of lipid-modifying therapies through drug target mendelian randomisation. PLoS biology, 20(2), e3001547. 10.1371/journal.pbio.3001547

[12] Malik, R., Chauhan, G., Traylor, M., Sargurupremraj, M., Okada, Y., Mishra, A., Rutten-Jacobs, L., Giese, A. K., van der Laan, S. W., Gretarsdottir, S., Anderson, C. D., Chong, M., Adams, H. H. H., Ago, T., Almgren, P., Amouyel, P., Ay, H., Bartz, T. M., Benavente, O. R., Bevan, S., … Dichgans, M. (2018). Multiancestry genome-wide association study of 520,000 subjects identifies 32 loci associated with stroke and stroke subtypes. Nature genetics, 50(4), 524–537. 10.1038/s41588-018-0058-3

[13] Dominiczak, M. H., & Caslake, M. J. (2011). Apolipoproteins: metabolic role and clinical biochemistry applications. Annals of clinical biochemistry, 48(Pt 6), 498–515. 10.1258/acb.2011.011111

[14] Olofsson, S. O., & Borèn, J. (2005). Apolipoprotein B: a clinically important apolipoprotein which assembles atherogenic lipoproteins and promotes the development of atherosclerosis. Journal of internal medicine, 258(5), 395–410. 10.1111/j.1365-2796.2005.01556.x

[15] Badimon, L., & Vilahur, G. (2014). Thrombosis formation on atherosclerotic lesions and plaque rupture. Journal of internal medicine, 276(6), 618–632. 10.1111/joim.12296

[16] Hsu, M. C., Chang, C. S., Lee, K. T., Sun, H. Y., Tsai, Y. S., Kuo, P. H., Young, K. C., & Wu, C. H. (2013). Central obesity in males affected by a dyslipidemia-associated genetic polymorphism on APOA1/C3/A4/A5 gene cluster. Nutrition & diabetes, 3(3), e61. 10.1038/nutd.2013.2

[17] Zhao, G. J., Yin, K., Fu, Y. C., & Tang, C. K. (2012). The interaction of ApoA-I and ABCA1 triggers signal transduction pathways to mediate efflux of cellular lipids. *Molecular medicine (Cambridge*, Mass.), 18(1), 149–158. 10.2119/molmed.2011.00183

[18] Wang, Q., Dai, H., Hou, T., Hou, Y., Wang, T., Lin, H., Zhao, Z., Li, M., Zheng, R., Wang, S., Lu, J., Xu, Y., Liu, R., Ning, G., Wang, W., Bi, Y., Zheng, J., & Xu, M. (2023). Dissecting Causal Relationships Between Gut Microbiota, Blood Metabolites, and Stroke: A Mendelian Randomization Study. Journal of stroke, 25(3), 350–360. 10.5853/jos.2023.00381

[19] Jamka, M., Morawska, A., Krzyżanowska-Jankowska, P., Bajerska, J., Przysławski, J., Walkowiak, J., & Lisowska, A. (2021). Comparison of the Effect of Amaranth Oil vs. Rapeseed Oil on Selected Atherosclerosis Markers in Overweight and Obese Subjects: A Randomized Double-Blind Cross-Over Trial. International journal of environmental research and public health, 18(16), 8540. 10.3390/ijerph18168540

[20] Yusuf, S., Hawken, S., Ounpuu, S., Dans, T., Avezum, A., Lanas, F., McQueen, M., Budaj, A., Pais, P., Varigos, J., Lisheng, L., & INTERHEART Study Investigators (2004). Effect of potentially modifiable risk factors associated with myocardial infarction in 52 countries (the INTERHEART study): case-control study. Lancet (London, England), 364(9438), 937–952. 10.1016/S0140-6736(04)17018-9

[21] O’Donnell, M. J., Chin, S. L., Rangarajan, S., Xavier, D., Liu, L., Zhang, H., Rao-Melacini, P., Zhang, X., Pais, P., Agapay, S., Lopez-Jaramillo, P., Damasceno, A., Langhorne, P., McQueen, M. J., Rosengren, A., Dehghan, M., Hankey, G. J., Dans, A. L., Elsayed, A., Avezum, A., … INTERSTROKE investigators (2016). Global and regional effects of potentially modifiable risk factors associated with acute stroke in 32 countries (INTERSTROKE): a case-control study. Lancet (London, England), 388(10046), 761–775. 10.1016/S0140-6736(16)30506-2

[22] Jung, C. H., Hwang, J. Y., Shin, M. S., Yu, J. H., Kim, E. H., Bae, S. J., Yang, D. H., Kang, J. W., Park, J. Y., Kim, H. K., & Lee, W. J. (2013). Association of apolipoprotein b/apolipoprotein A1 ratio and coronary artery stenosis and plaques detected by multi-detector computed tomography in healthy population. Journal of Korean medical science, 28(5), 709–716. 10.3346/jkms.2013.28.5.709

[23] Khan, M., Wasay, M., O’Donnell, M. J., Iqbal, R., Langhorne, P., Rosengren, A., Damasceno, A., Oguz, A., Lanas, F., Pogosova, N., Alhussain, F., Oveisgharan, S., Czlonkowska, A., Ryglewicz, D., & Yusuf, S. (2023). Risk Factors for Stroke in the Young (18-45 Years): A Case-Control Analysis of INTERSTROKE Data from 32 Countries. Neuroepidemiology, 57(5), 275–283. 10.1159/000530675

[24] Chen, S., Yang, F., Xu, T., Wang, Y., Zhang, K., Fu, G., & Zhang, W. (2021). Appraising the Causal Association of Plasma Homocysteine Levels With Atrial Fibrillation Risk: A Two-Sample Mendelian Randomization Study. Frontiers in genetics, 12, 619536. 10.3389/fgene.2021.619536

[25] Park, S., Lee, S., Kim, Y., Cho, S., Kim, K., Kim, Y. C., Han, S. S., Lee, H., Lee, J. P., Joo, K. W., Lim, C. S., Kim, Y. S., & Kim, D. K. (2021). Causal Effects of Homocysteine, Folate, and Cobalamin on Kidney Function: A Mendelian Randomization Study. Nutrients, 13(3), 906. 10.3390/nu13030906

